# OPTIMIZATION OF PERIOPERATIVE ANTIBIOTIC PROPHYLAXIS IN ONCOUROLOGY: THE ROLE OF A CLINICAL PHARMACOLOGIST AND ASSESSMENT OF CLINICAL AND ECONOMIC OUTCOMES

**DOI:** 10.64898/2026.04.07.26350335

**Authors:** Narine M. Dovlatbekyan, Irina N. Ochakovskaya, Artyom G. Penjoyan, Albert D. Avagimov, Vladimir M. Durleshter, Vladimir V. Onopriev

**Affiliations:** Kuban State Medical University, Ministry of Health of the Russian Federation; 4 Sedina St., Krasnodar, 350063, Russia; Regional Clinical Hospital No. 2, Ministry of Health of Krasnodar Krai; 6/2 Krasnykh Partizan St., Krasnodar, 350012, Russia

**Keywords:** antibiotic prophylaxis, antimicrobial stewardship, drug resistance, clinical pharmacologist, cost-benefit analysis, oncourology

## Abstract

**Objective:** To evaluate the effectiveness of a bundle of interventions involving a clinical pharmacologist aimed at changing surgeons’ approach to perioperative antibiotic prophylaxis (PAP) in an oncourology department.

**Materials and Methods:** A single-center retrospective observational study was conducted. Data from 226 patients who underwent prostatectomy or nephrectomy in the oncourology department of Regional Clinical Hospital No. 2 (Krasnodar, Russia) between 2023 and 2025 were analyzed. Periods before (n=125) and after (n=101) the implementation of an Antimicrobial Stewardship (AMS) strategy bundle with active participation of a clinical pharmacologist (pre-authorization, audit with feedback, education, “handshake stewardship”) were compared. The primary endpoint was the proportion of surgeries performed in compliance with the PAP protocol. Secondary endpoints included the incidence of infectious complications, antibiotic consumption (DDD/100 bed-days), direct costs of antibacterial drugs, dynamics of the microbial landscape, and the Drug Resistance Index (DRI).

**Results:** After AMS implementation, the proportion of surgeries performed in accordance with the PAP protocol increased from 0% to 47.7% for prostatectomies and to 55.6% for nephrectomies. The mean duration of antibiotic use decreased from 7 to 2 days (p<0.001). Antibiotic consumption decreased by 31.2%, and costs were reduced by a factor of 4.3. The proportion of ESKAPE organisms in the department’s microbial profile decreased from 26.3% to 16.4%. There was no statistically significant increase in the frequency of infectious complications (2.4% vs. 3.0%; p=1.000) or mortality (0% in both groups).

**Conclusions:** AMS implementation integrating a clinical pharmacologist into the oncourology department workflow significantly improved adherence to clinical guidelines, reduced irrational antibiotic use and financial costs without compromising patient safety. This approach can serve as a model for optimizing PAP in other surgical departments.

## Introduction

Antimicrobial resistance has been recognized by the World Health Organization as one of the leading global threats to modern healthcare [1]. Irrational prescribing of antibacterial drugs (ABDs), including excessively prolonged perioperative antibiotic prophylaxis (PAP), contributes to the selection of resistant strains and increased healthcare costs [2–4].

Despite international and Russian guidelines recommending that PAP be limited to a single preoperative antibiotic dose [5–9], surgeons commonly continue the traditional practice of prolonged PAP regimens [10–11], as the heavy responsibility for patient safety and physician fear regarding potential prognostic worsening prevail over rational prescribing [12]. This necessitates a comprehensive approach to optimizing antibiotic use with the involvement of clinical pharmacologists [13–15].

Implementation of Antimicrobial Stewardship (AMS) programs has traditionally focused on departments with high mortality [16–17]. However, effective antimicrobial resistance control requires extending stewardship strategies to all departments, including those with minimal mortality [14, 18]. In the oncourology department, where the risk of infectious complications remains low (particularly for “clean” surgeries such as prostatectomy and nephrectomy), the persistence of excessive antibiotic prescribing not only increases the financial burden on the healthcare institution but also promotes the selection of resistant organisms and increases the risk of adverse drug reactions [19–21]. Oncourology patients are often subject to prolonged follow-up and repeated invasive procedures, and may require chemo- and radiation therapy, potentially increasing their susceptibility to infections [22]. Therefore, minimizing the risk of antibiotic resistance development in this patient population is critically important.

Evidence on the effectiveness of AMS implementation in low-mortality departments remains limited [23]. In particular, there is insufficient data on which interventions most effectively optimize antimicrobial therapy, and their impact on clinical outcomes, economic indicators, and the departmental microbial landscape.

This study presents the experience and results of optimizing antibiotic use by oncourology department physicians, achieved through the implementation of a comprehensive approach whose key element is the synergistic interaction of the clinical pharmacologist with the treating physician team.

### Objective

to evaluate the effectiveness of interventions involving a clinical pharmacologist aimed at changing surgeons’ approach to perioperative antibiotic prophylaxis in an oncourology department.

## Materials and Methods

### Study design

A single-center retrospective observational study.

### Population and selection criteria

The study included data from 226 patients who underwent prostatectomy or nephrectomy in the oncourology department of Regional Clinical Hospital No. 2 between June 1, 2023, and March 31, 2025.

### Inclusion criteria

patients with oncourological pathology who underwent elective prostatectomy or nephrectomy.

### Exclusion criteria

patients who underwent combined surgical procedures (e.g., prostatectomy + cholecystectomy, cystoprostatectomy); absence of necessary information on PAP and CRP dynamics in the medical records.

To assess comparability of groups, demographic data, comorbidities, type and duration of surgery were analyzed.

### Description of the intervention

Between April and June 2024, an AMS intervention bundle with active participation of a clinical pharmacologist was implemented. The intervention included two phases:

#### Phase 1 (2022)

Development and formal implementation of a local PAP protocol with detailed descriptions of surgical procedures and antibiotic prophylaxis regimens. General physician education was conducted.

#### Phase 2 (April–June 2024, active phase)

Integration of the clinical pharmacologist into the department workflow with the following interventions:

- **Pre-authorization:** Prescription of second- and third-line antibiotics only after approval by the clinical pharmacologist (20 min/day).
- **Regular audit and feedback:** Daily review of medical records with real-time oral and written recommendations to treating physicians (20 min/day).
- **Staff education:** Regular short educational sessions on rational antibiotic therapy and PAP (once monthly, 30 min).
- **Handshake stewardship:** Daily personal interaction between the clinical pharmacologist and surgeons for joint discussion and decision-making on antibiotic therapy strategy, positive reinforcement of correct prescribing, and psychological support (20 min/day per treating physician).

### Observation periods and groups

Pre-intervention period (control group): June 1, 2023 – March 31, 2024 (125 patients). Post-intervention period (study group): June 1, 2024 – March 31, 2025 (101 patients).

### Endpoints

Primary endpoint: the proportion of surgeries (prostatectomy, nephrectomy) performed in strict compliance with the approved PAP protocol (single preoperative antibiotic dose). Secondary endpoints: incidence of infectious complications (surgical site infections) and mortality; antibiotic consumption calculated as DDD per 100 bed-days; direct antibiotic costs per patient and for the department overall; dynamics of the microbial landscape (proportion of ESKAPE organisms) and Drug Resistance Index (DRI) for key uropathogens (Escherichia coli, Klebsiella pneumoniae, Enterococcus faecalis).

### Microbiological analysis

was based on routine bacteriological studies (urine culture, wound discharge culture) within standard clinical patient management. The Drug Resistance Index (DRI) was calculated for key uropathogens as the product of the organism’s resistance rate to an antibiotic (as a fraction) and the consumption of that antibiotic in the department (DDD/100 bed-days), summed across all antibiotics potentially active against the given pathogen.

### Statistical analysis

Statistical analysis was performed using StatTech v. 4.8.11. Quantitative data were tested for normality using the Kolmogorov–Smirnov test. For normally distributed variables (CRP levels), the Student’s t-test for independent samples was used. Non-normally distributed data were described using the median (Me) and interquartile range (Q1–Q3). Categorical data were described as absolute values and percentages. Group comparisons for quantitative variables were performed using the Mann–Whitney U test; for categorical variables, the χ^2^ test or Fisher’s exact test was used. Differences were considered statistically significant at p<0.05. Sample size was not calculated a priori.

## Results

### Patient Characteristics

A total of 226 medical records were analyzed. The groups were comparable in sex, age, comorbidities (obesity, diabetes mellitus, chronic kidney disease), type and mean duration of surgery (Table 1). Comparability of groups regarding cancer staging and preoperative treatment volume was not assessed.

**Table 1.** Clinical characteristics of patients.

| Characteristic | Group 1 (control), n=125 | Group 2 (study), n=101 | p-value |
| --- | --- | --- | --- |
| Sex, male, n (%) | 109 (87.2%) | 85 (84.2%) | 0.514 |
| Sex, female, n (%) | 16 (12.8%) | 16 (15.8%) |  |
| Age, years, Me [Q1; Q3] | 65 [62; 70] | 64 [61; 68] | 0.125 |
| Obesity, n (%) | 46 (36.8%) | 37 (36.6%) | 0.979 |
| Diabetes mellitus, n (%) | 18 (14.4%) | 10 (9.9%) | 0.307 |
| Chronic kidney disease, n (%) | 1 (0.8%) | 0 (0.0%) | 1.000 |
| Surgery duration, min, Me [Q1; Q3] | 150 [130; 180] | 140 [120; 170] | 0.128 |
| Prostatectomy, n (%) | 89 (71.2%) | 65 (64.4%) | 0.272 |
| Nephrectomy, n (%) | 36 (28.8%) | 36 (35.6%) |  |

AMS implementation significantly increased the proportion of surgeries performed in strict compliance with the PAP protocol (Table 2).

**Table 2.** Adherence to the PAP protocol.

| Type of surgery | Pre-intervention period | Post-intervention period |
| --- | --- | --- |
| Prostatectomy | 0 / 89 (0%) | 31 / 65 (47.7%) |
| Nephrectomy | 0 / 36 (0%) | 20 / 36 (55.6%) |

### Clinical Outcomes

The incidence of infectious complications did not differ significantly between the study and control groups (2.4% vs. 3.0%; p=1.000). There was no mortality in either group.

The postoperative CRP increase, commonly observed in the early postoperative period and representing the main reason for extending antibiotic prophylaxis or switching to reserve antibiotics, was present in both groups (Table 3). CRP levels on postoperative days 3–4 were significantly higher in the study group (110.6 ± 75.9 mg/L vs. 75.3 ± 53.3 mg/L; p=0.012); however, by days 6–7, the differences resolved (43.7 ± 47.5 mg/L vs. 32.3 ± 39.1 mg/L; p=0.243). CRP distribution did not differ from normal (Kolmogorov–Smirnov test, p>0.05).

**Table 3.** Trends in CRP levels and infectious complications.

| Parameter | Control group (n=125) | Study group (n=101) | p-value |
| --- | --- | --- | --- |
| Rate of infectious complications, % (n) | 2.4% (3) | 3.0% (3) | 1.000 |
| CRP on days 3–4, mg/L (M ± SD) | 75.3 ± 53.3 | 110.6 ± 75.9 | 0.012 |
| CRP on days 6–7, mg/L (M ± SD) | 32.3 ± 39.1 | 43.7 ± 47.5 | 0.243 |

CRP — C-reactive protein

**Figure 1.**
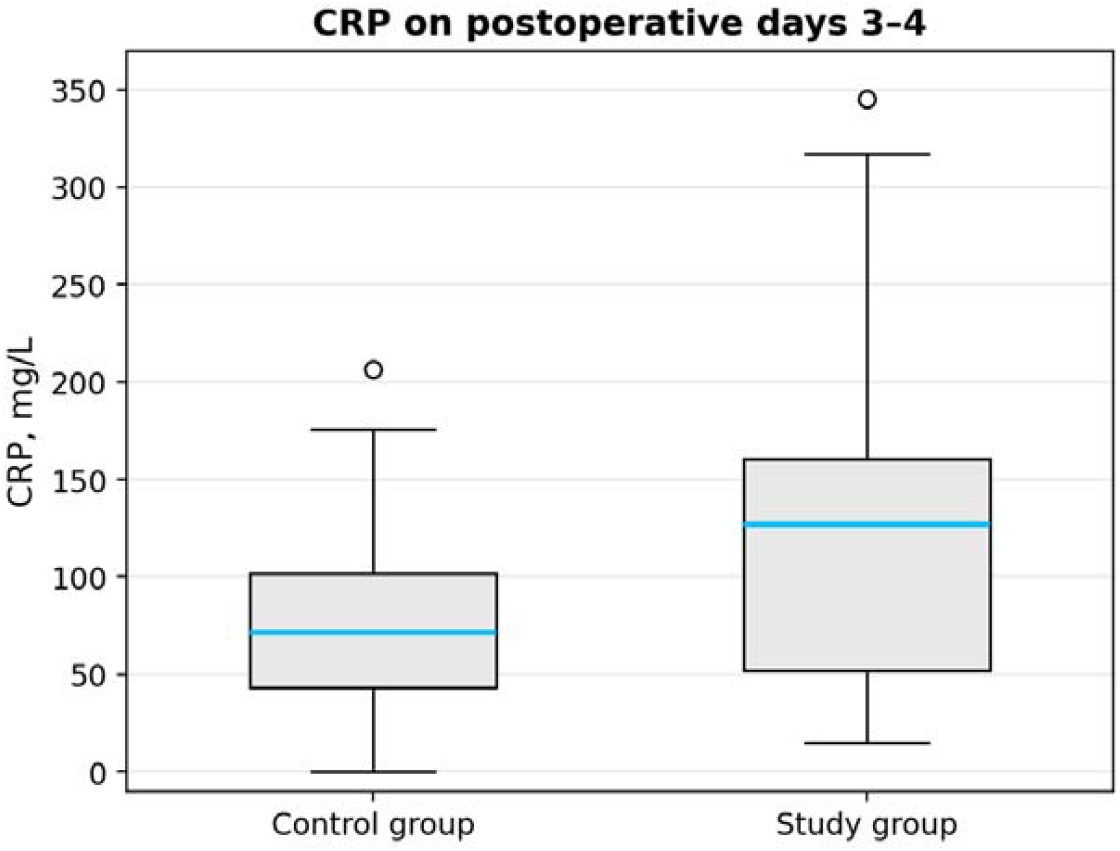
CRP levels on postoperative days 3–4 (box plot).

### Pharmacoepidemiological and Pharmacoeconomic Results

AMS implementation led to a statistically significant reduction in the mean duration of antibiotic use from 7 to 2 days (p<0.001) and a 3.4-fold reduction in median antibiotic cost per patient (from 298.40 RUB to 87.80 RUB; p<0.001) (Table 4). Direct antibiotic costs for the department decreased by a factor of 4.3 (from 1,187,308 RUB to 275,815 RUB). Overall antibiotic consumption decreased by 31.2% (from 44.6 to 30.7 DDD/100 bed-days). There was a substantial reduction in the consumption of reserve antibiotics, particularly carbapenems (from 3.7 to 0.6 DDD/100 bed-days). Cephalosporin consumption increased as a result of greater physician adherence to the PAP protocol, updated in accordance with current clinical guidelines [7].

**Table 4.**
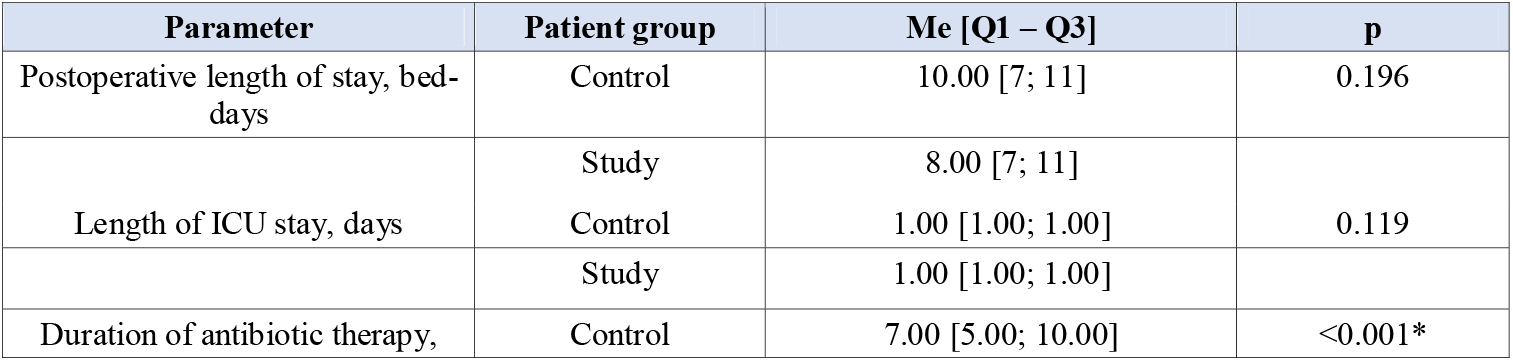
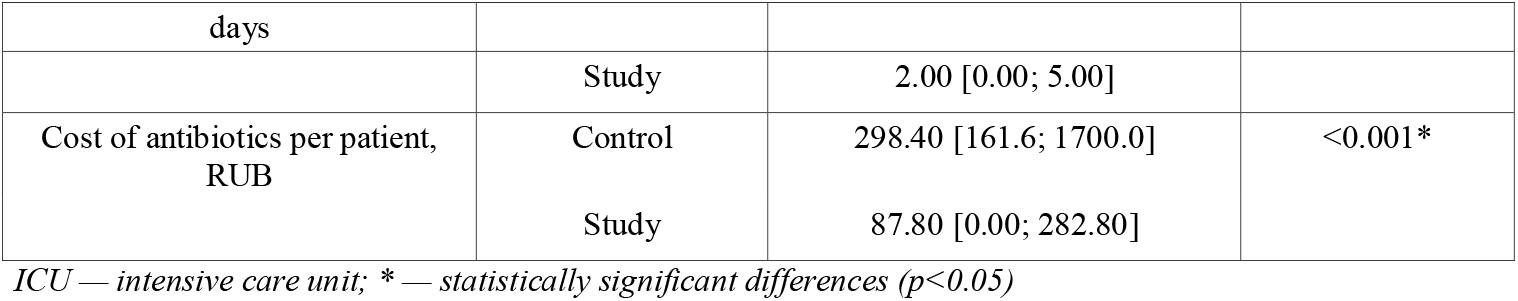
Pharmacoepidemiological and economic indicators.

| Parameter | Patient group | Me [Q1 – Q3] | p |
| --- | --- | --- | --- |
| Postoperative length of stay, bed-days | Control | 10.00 [7; 11] | 0.196 |
|  | Study | 8.00 [7; 11] |  |
| Length of ICU stay, days | Control | 1.00 [1.00; 1.00] | 0.119 |
|  | Study | 1.00 [1.00; 1.00] |  |
| Duration of antibiotic therapy, | Control | 7.00 [5.00; 10.00] | <0.001* |
| days |  |  |  |
|  | Study | 2.00 [0.00; 5.00] |  |
| Cost of antibiotics per patient, RUB | Control | 298.40 [161.6; 1700.0] | <0.001* |
|  | Study | 87.80 [0.00; 282.80] |  |

ICU — intensive care unit; \* — statistically significant differences ( $p < 0.05$ )

**Figure 2.**
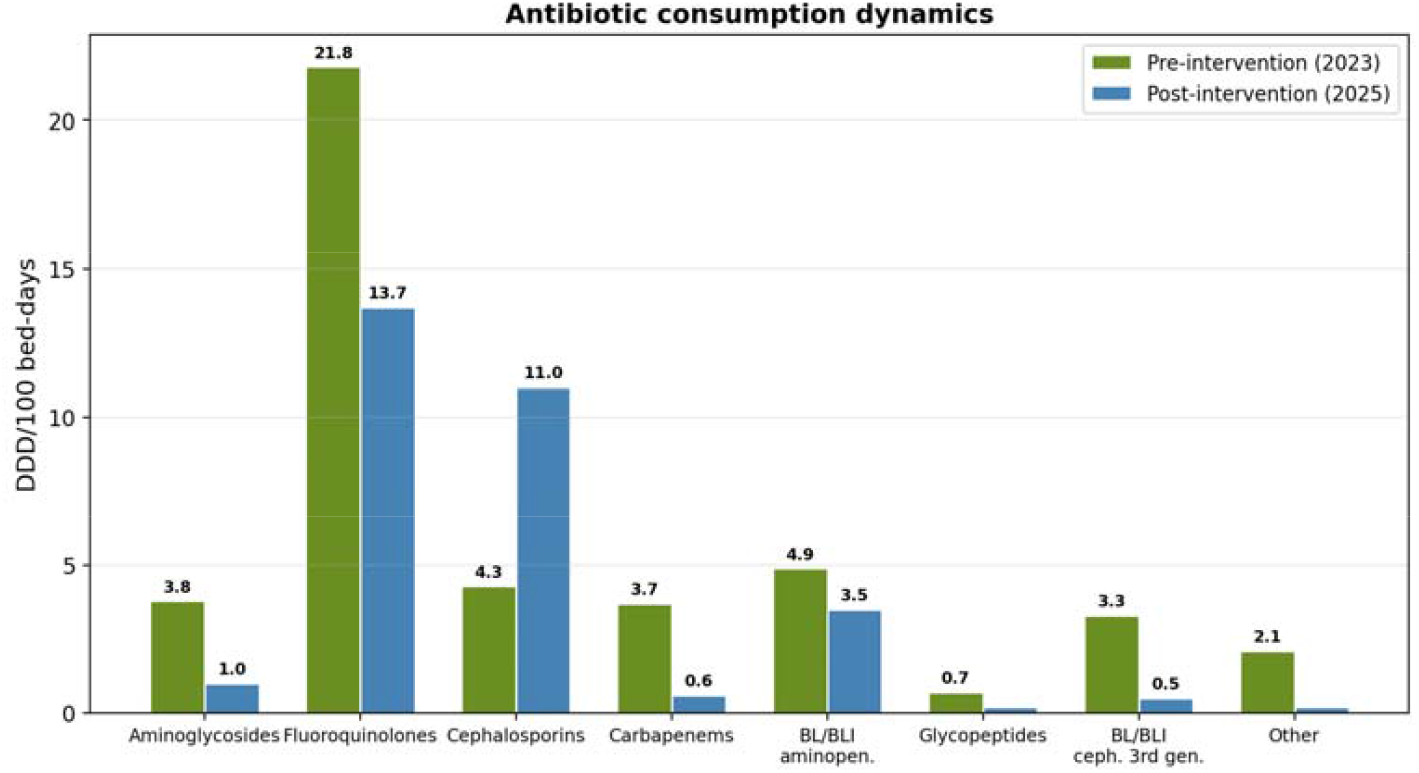
Changes in antibiotic consumption (DDD/100 bed-days) between the pre- and post-intervention periods.

### Microbiological Results

During the observation period, the proportion of ESKAPE organisms (Enterococcus faecium, Staphylococcus aureus, Klebsiella pneumoniae, Acinetobacter baumannii, Pseudomonas aeruginosa, Enterobacter spp.) in the overall departmental microbial profile decreased from 26.3% to 16.4%. Notably, the isolation rate of K. pneumoniae decreased from 14.1% to 4.7%. Analysis of the Drug Resistance Index (DRI) revealed divergent trends: a decrease in DRI for E. faecalis and K. pneumoniae was achieved, while an increase in DRI for E. coli was observed (0.72 vs. 0.51). The increase in DRI for E. coli during the post-intervention period was primarily driven by increased resistance rates to third-generation cephalosporins (from 55.9% to 80.6%) and fluoroquinolones (from 73.5% to 83.9%). No statistically significant linear correlation was found between total cephalosporin consumption in DDD/100 bed-days and DRI dynamics for E. coli over the entire observation period (r = –0.12; p>0.05). This may indicate the complex nature of resistance development, including the possible influence of horizontal gene transfer and the delayed effect of selective pressure.

**Figure 3.**
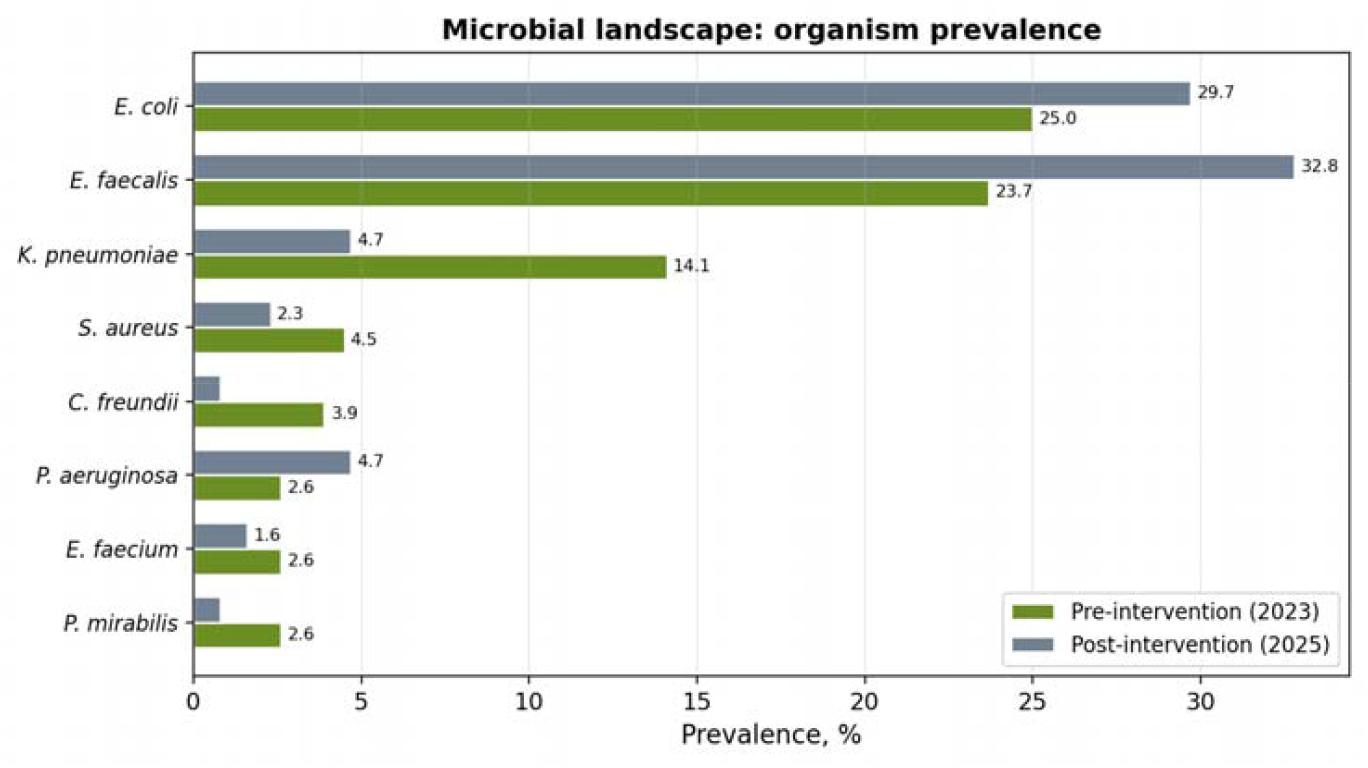
Trends in the prevalence of ESKAPE organisms within the department’s microbial profile.

**Figure 4.**
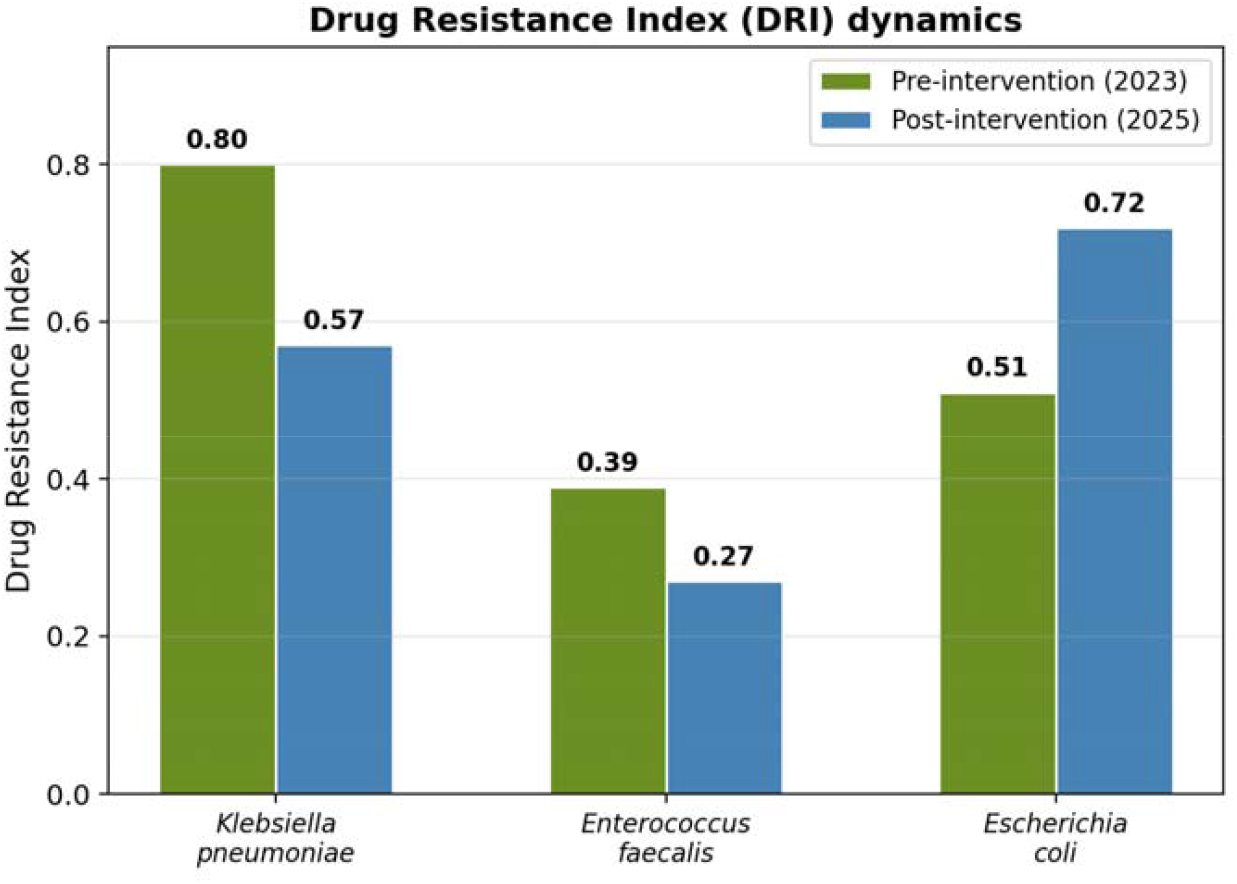
Dynamics of the Drug Resistance Index (DRI).

## Discussion

The present study demonstrates that implementing a multicomponent AMS program with integration of a clinical pharmacologist into the oncourology department workflow achieved significant improvements across multiple domains. The main result was a substantial increase in PAP protocol adherence — from 0% to 47.7% for prostatectomy and 55.6% for nephrectomy, indicating the effectiveness of the applied approach in changing established surgical practice.

Although the intervention bundle included several components, we hypothesize that the personalized “handshake stewardship” approach played the key role in changing physician behavior. Informal communication, shared decision-making, and positive reinforcement helped overcome the psychological barrier — the fear of potential infectious complications when reducing PAP duration — which traditionally dominates over rational antibiotic prescribing [13–15].

The achieved results are consistent with data from international studies confirming the effectiveness of AMS programs [13, 19–21, 23–24]. In particular, our study extends the findings of Gebretekle et al. [24], demonstrating that active clinical pharmacologist involvement is effective not only in high-mortality departments but also in the oncourology setting with inherently low infectious complication risk. Furthermore, we identified not only reduced antibiotic consumption but also a substantial direct economic effect — a 4.3-fold reduction in costs.

Our findings are consistent with data from Karpov et al. [23], who also confirmed the effectiveness of AMS implementation in a Russian hospital setting. While their work demonstrates a comprehensive hospital-wide effect, our study identified the advantages of a targeted approach in oncourology, where clinical pharmacologist integration enabled both rapid, significant improvement in PAP protocol adherence without increased infectious complication risk and substantial reduction in antibiotic therapy costs. This confirms the value of specialized antimicrobial programs in individual surgical profiles from both clinical and economic perspectives.

An important aspect of our study is the comprehensive assessment of the intervention’s impact on the microbial landscape and antibiotic resistance. We found divergent trends: a decrease in the proportion of ESKAPE organisms from 26.3% to 16.4% and a reduction in DRI for K. pneumoniae and E. faecalis, consistent with reduced use of carbapenems and aminoglycosides. However, the increase in E. coli resistance to cephalosporins and fluoroquinolones, despite reduced consumption of the latter, points to the complex nature of resistance formation. This phenomenon may be explained by both selective pressure from increased use of third-generation cephalosporins for PAP and horizontal transfer of resistance genes, warranting further investigation and underscoring the need for continuous microbiological monitoring even against the background of successful AMS implementation.

An interesting finding was the statistically significant elevation of CRP on days 3–4 in the study group, which, however, did not lead to an increase in infectious complications and resolved by days 6–7. This may reflect a more pronounced but controlled systemic inflammatory response in patients not receiving prolonged PAP and requires further investigation to determine the clinical significance of this dynamic.

## Limitations

Study limitations include its retrospective design, single-center nature, and relatively short observation period. Sample size was not calculated a priori, which may have limited the ability to detect statistically significant differences for some secondary endpoints (e.g., infectious complication rate). We did not analyze data on cancer staging, tumor histological characteristics, or preoperative chemo- or radiation therapy, which could have affected group comparability regarding infectious complication risk. The influence of other potential confounders (changes in surgical technique or infection control protocols during the study period) was also not assessed. Furthermore, restricting the sample to prostatectomy and nephrectomy patients limits generalizability to all oncourological procedures.

## Conclusions

Implementation of AMS interventions in the oncourology department with active clinical pharmacologist involvement significantly improved PAP protocol adherence, reduced irrational antibiotic use and financial costs, and had a positive impact on the microbial landscape without increasing the incidence of infectious complications. The presented model, based on interdisciplinary collaboration and personalized feedback, is an effective and low-cost tool that can be successfully scaled to other surgical departments for optimizing PAP and combating the growth of antimicrobial resistance.

## Declarations

### Ethics statement

This study was approved by the Ethics Committee of the State Budgetary Healthcare Institution “Regional Clinical Hospital No. 2” of the Ministry of Health of Krasnodar Krai (Krasnodar, Russia), meeting No. 110, dated October 16, 2024. Due to the retrospective nature of the study, written informed patient consent was not required.

### Competing interests

The authors declare no competing interests.

### Funding

This research received no specific grant from any funding agency in the public, commercial, or not-for-profit sectors.

### Data availability

The data supporting the findings of this study are available from the corresponding author upon reasonable request.

### Author contributions

All authors contributed equally to all stages of the work and preparation of this article.

## Acknowledgments

The authors express their gratitude to the medical staff of the oncourology department of Regional Clinical Hospital No. 2 for their cooperation and assistance in conducting the study.

## Author Information

Narine M. Dovlatbekyan — Department of Clinical Pharmacology and Functional Diagnostics, Kuban State Medical University; Clinical Pharmacologist, Regional Clinical Hospital No. 2.

Irina N. Ochakovskaya — Department of Clinical Pharmacology and Functional Diagnostics, Kuban State Medical University; Head of the Clinical Pharmacology Department, Regional Clinical Hospital No. 2.

Artyom G. Penjoyan — Department No. 2, Regional Clinical Hospital No. 2.

Albert D. Avagimov — Oncology Department No. 2, Regional Clinical Hospital No. 2.

Vladimir M. Durleshter — Deputy Chief Physician for Surgery, Regional Clinical Hospital No. 2; Head of the Department of Surgery No. 3, Kuban State Medical University.

Vladimir V. Onopriev — Head of the Department of Clinical Pharmacology and Functional Diagnostics, Kuban State Medical University.

## Notes

### Competing Interest Statement

The authors have declared no competing interest.

### Author Declarations

This study was approved by the Ethics Committee of the State Budgetary Healthcare Institution Regional Clinical Hospital No. 2 of the Ministry of Health of Krasnodar Krai (Krasnodar, Russia), meeting No. 110, dated October 16, 2024. Due to the retrospective nature of the study, written informed patient consent was not required

